# High prevalence of hypertension and high-normal blood pressure: findings from a large population-based survey of young adults in Zimbabwe

**DOI:** 10.1101/2023.07.03.23292156

**Authors:** Kalpana Sabapathy, Fredrick Cyprian Mwita, Ethel Dauya, Tsitsi Bandason, Victoria Simms, Chido Dziva Chikwari, Aoife M Doyle, David Ross, Anoop Shah, Richard Hayes, Aletta E Schutte, Katharina Kranzer, Rashida Abbas Ferrand

## Abstract

**Background:** Most cardiovascular mortality is due to hypertension and onset may be in youth. We investigated the prevalence of and risk factors for elevated blood pressure (BP) (hypertension (≥140/90mmHg) and high-normal BP (130-139/85-89mmHg)) among youth in Zimbabwe.

**Methods:** A population-based survey of randomly sampled 18-24 year olds from 24 communities in three provinces was conducted, with standardised questionnaires to collect socio-demographic, behavioural and clinical data. Height, weight and BP were recorded. The association of potential risk factors with elevated BP was examined using multivariable logistic regression.

**Findings:** Of 17,682 participants recruited (98% of those eligible), 17,637 had complete data. The median age was 20 (IQR: 19-22) years and 60.7% were female. After excluding pregnant women (N=754), the prevalence of hypertension and high-normal BP was 7.4% (95% CI:7.0-7.8) and 12.2% (95% CI:12.7-13.7), respectively. Prevalence of hypertension was higher in men (8.7% (95% CI:8.2-9.6) vs 6.6% (95% CI:6.0-6.9) in men and women, respectively) but with age increased to equivalent levels among women (at 18y 7.3% (95% CI:6.2-8.6) and 4.3% (95% CI:3.5-5.2); at 23-24 years 10.9% (95% CI:9.3-12.5) and 9.50% (95% CI:8.4-10.7) in men and women, respectively). After adjusting for confounders, male sex ((aOR) 1.53 (95% CI:1.36-1.74)), increasing age (19-20 years aOR:1.20 (1.00-1.44); 21-22 years aOR:1.45 (95% CI 1.20-1.75); 23-24 years aOR 1.90 (95% CI:1.57-2.30), vs 18 years) and obesity (aOR 1.94 (95% CI:1.53-2.47)) were associated with hypertension. Being underweight (aOR 0.79 (0.63-0.98)) and living with HIV (aOR 0.71 (95% CI:0.55-0.92)) were associated with lower odds of hypertension.

**Interpretation:** Prevalence of elevated BP is high among youth in SSA and rises rapidly with age. Further research is needed to understand drivers of BP elevation and the extent of target organ damage in youth in SSA, to guide implementation of prevention and management strategies.

**Funding:** Wellcome Trust.

## Background

Worldwide, 1.3 billion adults aged 30-79 years have hypertension and it is the commonest cause of cardiovascular disease.^1^ Raised systolic blood pressure is the leading cause of death globally, causing more than 10.8 million deaths (19% of all deaths) in 2019, and contributing to 9% of diasability-adjusted life years lost.^2^

Low and middle-income countries (LMICs) have experienced a progressive rise in the number of adults with hypertension surpassing that of high-income countries (HICs), with an estimated 1.04 billion people living with hypertension in LMICs compared to 349 million in HICs.^2,3^ Where hypertension was a condition associated with affluence, it is now one of poverty.^4^ Sub-Saharan Africa (SSA) has been undergoing a rapid epidemiological transition and the prevalence and incidence of hypertension are increasing.^5^ The World Health Organization (WHO) estimates that Africa has the highest prevalence of hypertension and the highest age-adjusted rates of cardiovascular disease of any global region.^2^

Hypertension was understood to be a disease of advancing age. Existing international screening and treatment guidelines on hypertension are predominantly derived from studies in older adults with an average age of about 50 years and in people of European ancestry.^6-8^ Incidence of hypertension in individuals of African origin occurs earlier and blood pressure (BP) elevations are more severe than in those of European ancestry.^5^ Population surveys in SSA among individuals with average age of 30-50 years have shown a high prevalence of hypertension.^9-11^ However, evidence is accumulating that hypertension is prevalent even among adolescents and young people in SSA with an estimated 6-10% having BP that meet criteria of hypertertion. In addtion, in one study, about a third of adolescents and young people had an elevated BP that did not meet the criteria for hypertension.^12,13^ Elevated BP frequently progresses to hypertension and is an independent risk factor for cardiovascular disease, and elevations in youth track into adulthood.^14-16^ These studies have had small sample sizes, often recruited adolescents and young people as part of a larger group of adults and not assessed age-specific risk factor or been conducted in schools in settings where school attendance is not universal .^12,17^

There is scant understanding of the epidemiology of hypertension in young people, and especially so in SSA.^5,18^ Using data from a large population-representative survey in Zimbabwe, we aimed to investigate the population distribution of blood pressure and the prevalence of hypertension (≥140 and/or 90mmHg) including isolated systolic and diastolic hypertension, and high-normal BP (130-139 and/or 85-89mmHg)^6^, and factors associated with hypertension among young adults in Zimbabwe.

## Methods

### Study design and setting

A population-based survey was conducted in Zimbabwe among 18-24 year-olds to ascertain the outcome of a cluster-randomised trial (CHIEDZA) investigating the impact of community-based integrated HIV and sexual and reproductive health services for youth on population-level HIV outcomes (Trial registration number NCT03719521). The trial protocol, including survey methods has been published.^19^ Briefly, the trial was conducted in 24 urban and peri-urban communities in three provinces (Harare, Bulawayo, and Mashonaland East), with eight communities per province. Each community had geographically demaracated areas that served as clusters. The 24 clusters were randomised 1:1 to either standard of care (existing facility-based health services) or the intervention, stratified by province. Implementation of the trial outcome cross-sectional survey was staggered with recruitment in Harare from October to December 2021, in Bulawayo from January to March 2022 and in Mashonaland East from April to June 2022, aiming to recruit a total of 16,800 participants (5,600 per province).

### Survey methods

Multi-stage sampling was used. Satellite images were utilised to map each building within a cluster onto OpenStreetMap, and ARCGIS was utilized to divide all streets within the cluster into short sections (approximately 100-200m). A random sample of street sections was selected, and all residents of those sections were enumerated. All eligible individuals (aged 18-24 years) residing in households on either side of the selected section were approached for participation in the survey.

An interviewer-adminstered questionnaire was used to collect socio-demographic and behavioural data and medical history including pregnancy, knowledge of HIV status, history of chronic conditions, use of regular medication and self-rated health. The International Physical Activity Questionnaire was used to ascertain levels of physical activity.^20^ The Alcohol Use Disorders Identification Test (AUDIT), a 10-item intenationally validated tool, was used to screen for alcohol use disorder.^21^ The Shona Symptom Questionnaire (SSQ), a locally developed and validated 14-item scale, was used to screen for common mental disorders.^22^

Weight was measured to the nearest 0.1 kg using digital Seca^®^ 803 weight scales (Seca, Hamburg, Germany) in minimal clothing and shoes removed. Height was measured to the nearest 0.1 centimeter using a Seca^®^ 213 stadiometer (Seca). Three seated BP measurements were taken at standardised time points during interview (at least 5 minutes apart) using a digital sphygmomanometer (Omron X2, Kyoto, Japan), with the first measure taken after 15 minutes of rest. BP measurements were performed using cuffs sized on mid-uper arm circumference (17-22cm: small cuff; >22-32cm: regular cuff; >32-42cm: large cuff) according to WHO guidelines. A dried blood spot was collected for HIV antibody and viral load testing.

### Definitions

A principal component analysis of household assets was used to create a wealth index in quintiles. Levels of physical activity were expressed as multiples of the resting metabolic rate (MET) in minutes. A threshold of 8 on AUDIT score: low risk <8 and high risk ≥8 was used to define alcohol use disorder or hazardous drinking.^21^ A SSQ score of ≥8 indicated a risk of common mental disorders.^22^ Weight was categorised according to body mass index (BMI) as follows: underweight was BMI<18.5kg/m^2^; normal weight was a BMI 18.5–24.9kg/m^2^; overweight was a BMI 25.0–29.9 kg/m^2^ and obese was BMI ≥30.0 kg/m^2^. The mean of the second and third BP readings was used to determine BP outcomes.^7^ BP outcome categories were defined according to International Society of Hypertension (ISH) guidelines as follows: hypertension ≥140 and/or 90mmHg; isolated systolic hypertension as systolic BP ≥140mmHg (≤89mmHg diastolic BP); isolated diastolic hypertension as diastolic BP ≥90mmHg (≤139mmHg systolic BP); high-normal BP 130-139 and/or 85-89mmHg.^6^

### Data management and statistical analysis

Survey data were collected onto electronic tablets using the SurveyCTO (Cambridge, USA) platform and uploaded to a secure server at the end of each day, using a secure the Biomedical Research and Training Institute (BRTI) internet connection. Data were downloaded and stored in a password-protected database utilising Microsoft Access to execute quality control queries, with access limited to defined study personnel. Data were analysed using Stata v17.0 (StatCorp, College Station, TX, USA).

Participants who did not have three BP measurements were excluded from analysis (N=45). The prevalence and corresponding 95% confidence intervals of hypertension and high-normal BP by sex (and pregnancy status in women) were calculated. The distribution of systolic and diastolic BP in males and non-pregnant females were examined, and the prevalence of hypertension by age and obesity plotted in a two-way graph.

Logistic regression modelling was used to investigate factors associate with hypertension and high-nomal BP, adjusting for cluster. We excluded women who reported they were pregnant due to physiological differences in pregnancy which impact BP and potential to distort BMI data. Factors associated in univariate analysis at a significance level of p<0.05 were taken forward into a multivariable model. If co-linearity was plausible and supported by cross-tabulation of the data, only the variable which was more strongly associated with the outcome was retained in the models, and hence socio-economic quintiles were chosen over household income. Variables with a p-value <0.05 in the final model were regarded as independently associated with the outcome. Potential interactions between sex and age or sex and BMI for the association with hypertension were examined.

The sample size of the survey was determined by the underlying assumptions and power needed for detecting a difference in primary outcome by arm for the underlying trial for which the population-based survey was conducted.^19^

### Ethics

The study was approved by the Medical Research Council, Zimbabwe, the Biomedical Research and Training Institute Institutional Review Board and the ethics committee of the London School of Hygiene and Tropical Medicine. Participants viewed an information video about the study (in either English, Shona or Ndebele) on a tablet. Consent was documented electronically on a tablet, with participants retaining a signed paper copy for their records. Participants with a systolic BP

>180mmHg or/and diastrolic BP >100mmHg or a BP of 140/90mmHg with consistent symptoms were were referred to the nearest clinic for further management.

### Role of the funding source

The funder of the study had no role in study design, data collection, data analysis, data interpretation, or writing of the report. KS, TB, RAF and VS had full access to all data. KS had final responsibility for the decision to submit for publication.

## Results

Of the 18,043 eligible individuals identified by enumeration, 17,682 (98.0%) particpants were contactable, consented and were recruited. Among 17,637 particpants with three blood pressure measures the median age was 20 (IQR:19, 22) years and 60.7% were female. The majority of participants had completed secondary education up to Form 4 (i.e. 10 years of schooling) (61.0%), with 28.0% of all participants still in full-time education. Nearly half of all particpants were neither in education nor employed (Table 1).

**Table 1.** Participant characteristics.

|  | **Population**<br>**Overall (N=17,637)<sup>1</sup>** |
| --- | --- |
| ***Socio-demographic characteristics*** |  |
| **Sex** |  |
| Female | 10,727 (60.7%) |
| Male | 6,910 (39.2%) |
| **Age category (years)** |  |
| 18 | 4,204 (23.8%) |
| 19-20 | 5,022 (28.5%) |
| 21-22 | 4,127 (23.4%) |
| 23-24 | 4,284 (24.3%) |
| **Highest education attained** |  |
| Primary or below | 3,253 (18.4%) |
| Secondary form 4 | 10,761 (61.0%) |
| Secondary form 6 | 2,216 (12.6%) |
| Higher education | 1,407 (8.0%) |
| **Occupation** |  |
| In education | 4,943 (28.0%) |
| Employed/own business | 832 (4.7%) |
| Work in informal sector | 3,151 (17.9%) |
| None of the above | 8,711 (49.4%) |
| **Household income (per month)** |  |
| Less than US\$50 | 2,616 (14.8%) |
| US\$50-100 | 4,457 (25.3%) |
| US\$101-200 | 4,741 (26.9%) |
| US\$201-500 | 2,697 (15.3%) |
| >US\$500 | 593 (3.4%) |
| *Don't know/Missing* | *2,533 (14.4%)* |
| **Socio-economic quintile** |  |
| Lowest quintile (least affluent) | 4,029 (22.8%) |
| Second lowest quintile | 3,119 (17.7%) |
| Middle quintile | 3,539 (20.1%) |
| Second highest quintile | 3,437 (19.5%) |
| Highest quintile (most affluent) | 3,513 (19.9%) |
| ***Behavioural characteristics*** |  |
| **Physical activity (MET)<sup>2</sup>** |  |
| Low | 6,333 (35.9%) |
| Moderate | 5,765 (32.7%) |
| High | 5,321 (30.2%) |
| *Don't know/Missing* | *218 (1.2%)* |
| **Alcohol/risk of problem drinking** |  |
| Never drink/low risk alcohol | 16,763 (95.0%) |
| Increased risk - possible dependence (AUDIT score $\geq 8$)<sup>3</sup> | 816 (4.6%) |
| *Don't know/Missing* | *58 (0.33%)* |
| **Smoking history** |  |
| Never smoker | 16,571 (94.0%) |
| Ever smoker | 1055 (6.0%) |
| *Missing* | *11 (0.1%)* |
| ***Clinical characteristics*** |  |
| **Self-rated health in previous 12months** |  |
| Excellent | 7,220 (40.9%) |
| Good | 8,992 (51.0%) |
| Fair | 1,311 (7.4%) |
| Poor | 114 (0.7%) |
| **Known hypertension** |  |
| No | 17,502 (99.2%) |
| Yes | 133 (0.8%) |
| *Don't want to say* | *2 (0.01%)* |
| **Known diabetes** |  |
| No | 16,609 (99.8%) |
| Yes | 25 (0.1%) |
| *Don't want to say* | *3 (0.01%)* |
| **Known renal disease** |  |
| No | 16,616 (99.9%) |
| Yes | 18 (0.1%) |
| *Don't want to say* | *3 (0.02%)* |
| **Pregnancy history<sup>1</sup>** |  |
| Never pregnant | 6,029 (56.2%) |
| Previous pregnancy only | 3,903 (36.4%) |
| Currently pregnant | 754 (7.0%) |
| *Don't want to say* | 41 (0.4%) |
| **Shona symptom questionnaire** |  |
| Low risk of common mental disorder | 16,429 (93.2%) |
| Risk of common mental disorder (SSQ score $\geq 8$)<sup>4</sup> | 1,208 (6.9%) |
| **BMI category** |  |
| Underweight | 1,620 (9.2%) |
| Normal weight | 12,337 (69.8%) |
| Overweight | 2,884 (16.3%) |
| Obese | 795 (4.5%) |
| *Missing* | 1 (0.01%) |
| **HIV status** |  |
| HIV-negative | 16,291 (92.4%) |
| HIV-positive | 1,222 (6.9%) |
| *Missing/confirmatory results unavailable* | 124 (0.7%) |

1. For pregnancy N=10,272; 2. The International Physical Activity Questionnaire was used to ascertain levels of physical activity, expressed as multiples of the resting metabolic rate (MET) in MET minutes; 3. The Alcohol Use Disorders Identification Test (AUDIT), threshold of 8 on AUDIT score: low risk <8 and high risk $\geq 8$ ; 4 Standard SSQ thresholds of $\geq 8$ to indicate risk of common mental disorders was used vs <8 for low risk.

A third of participants self-reported low levels of physical activity. High risk alcohol use (AUDIT score ≥8) and ever smoking were reported by 4.6% and 6.0% of particpants respectively, the majority of whom were men (72.4% of alcohol drinkers and 89.9% of smokers). History of pregnancy was common with 36.4% of women reporting a previous pregnany and 7.0% of women were pregnant at the time of the survey. One-fifth of all particpants were overweight or obese and 9.2% were underweight (Table 1).

Self-rated health was excellent or good in 91.9% of individuals and only a small minority (<1%) reported a previous diagnosis of diabetes, chronic renal disease or hypertension. Of the 133 (0.8%) participants who were previously diagnosed with hypertension, 24 were on treatment and 17 had BP <140/90mmHg. The HIV prevalence was 6.9% and 32.0% (N=391) of participants living with HIV reported that they were taking antiretroviral therapy.

### Prevalence of hypertension and high-normal BP

The first blood pressure measurement was slightly higher than the subsequent two measures (Supplementary Table 1), supporting the use of the average of second and third measures to determine BP.

Excluding 754 self-reported pregnant women, the population median systolic and diastolic BP were 116 (IQR:110, 124) and 74 (IQR:69, 80) mmHg, respectively (Figure 1). The median systolic BP was higher among men (119 (IQR:112, 127) mmHg) than women (114 (IQR:108, 122) mmHg). The median diastolic pressure was lower among men (73 (IQR:69, 79) mmHg) than women (75 (IQR:70, 80) mmHg) (Table 2).

**Table 2:** Prevalence (with 95% confidence intervals) of hypertension, systolic and diastolic hypertension and high-normal BP.

|  | **Hypertension**<br>**≥140/90 mmHg** | **Systolic hypertension**<br>**≥140 mmHg/any diastolic** | **Isolated systolic**<br>**hypertension**<br>**≥140/≤89 mmHg** | **Diastolic hypertension**<br>**any systolic/≥90 mmHg** | **Isolated diastolic**<br>**hypertension**<br>**≤139 mmHg/≥90 mmHg** | **High-normal BP**<br>**130-139/85-89 mmHg** |
| --- | --- | --- | --- | --- | --- | --- |
| **Population overall<sup>1</sup>**<br>**(N=16,883)** | 1,254<br>7.4% (7.0, 7.8) | 698<br>4.1% (3.8, 4.4) | 542<br>3.2% (2.9, 3.5) | 712<br>4.3% (4.0, 4.6) | 556<br>3.4% (3.2, 3.7) | 2064<br>12.2% (12.7, 13.7) |
| **Non-pregnant women**<br>**(N=9,973)** | 642<br>6.6% (6.0, 6.9) | 290<br>2.8% (2.5, 3.1) | 187<br>1.9% (1.6, 2.2) | 455<br>4.6% (4.2, 5.0) | 366<br>3.7% (3.3, 4.1) | 1,033<br>10.4% (9.8, 11.0) |
| **Pregnant women**<br>**(N=754)** | 37<br>4.9% (3.5, 6.7) | 20<br>2.7% (1.6, 4.1) | 13<br>1.7% (0.9, 2.9) | 24<br>3.2% (2.0, 4.7) | 17<br>2.3% (1.3, 3.6) | 49<br>6.5% (4.8, 8.5) |
| **Men**<br>**(N=6910)** | 612<br>8.7% (8.2, 9.6) | 422<br>6.1% (5.6, 6.7) | 355<br>5.1% (4.6, 5.7) | 257<br>3.7% (3.3, 4.2) | 190<br>2.7% (2.4, 3.2) | 1,031<br>14.9% (14.1, 15.8) |

1. Exclude self-reported currently pregnant women (N=754)

**Figure 1:**
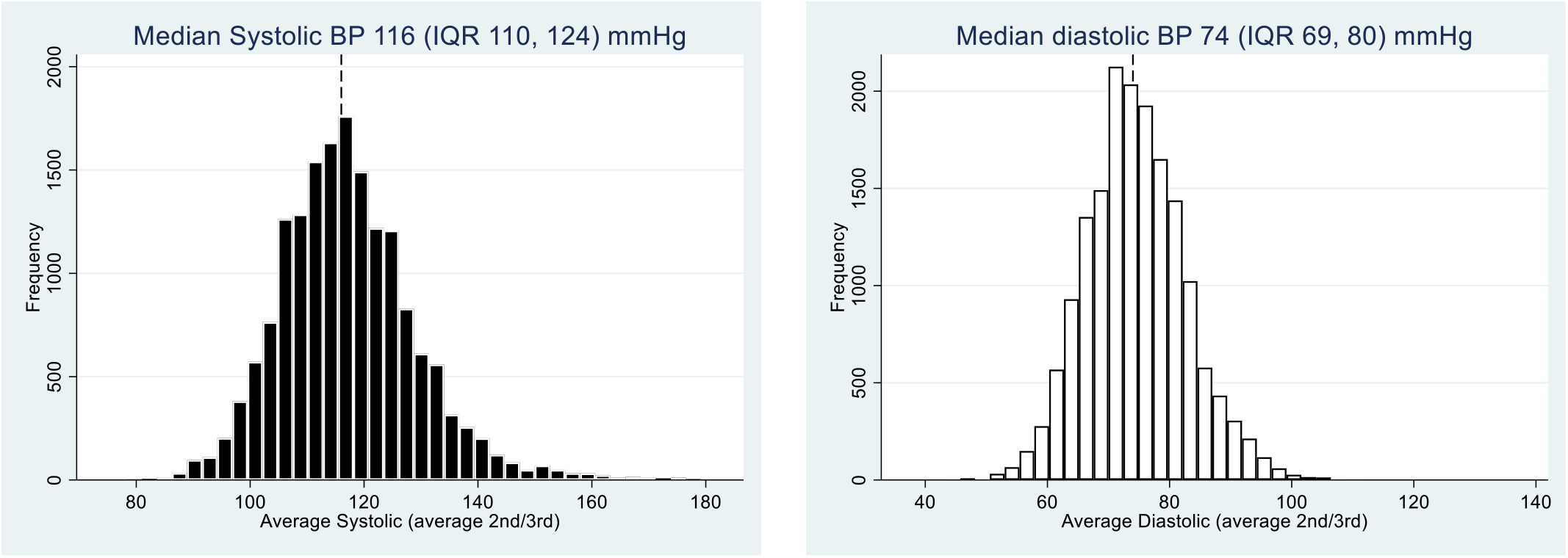
Population systolic and diastolic BP distribution (excluding pregnant women)

The prevalence of hypertension was 7.4% (95 %CI:7.0, 7.8), and 12.2% (95% CI:11.7, 12.7) had high-normal BP (Table 2). The prevalence of isolated systolic and diastolic hypertension were 3.2% (95% CI:2.9, 3.5) and 3.3% (95% CI:3.0, 3.6), respectively. Hypertension was more prevalent among men (8.7% (95% CI:8.2, 9.6) than women (6.6% (95% CI:6.0, 6.9) driven by higher prevalence of systolic BP elevation (Table 2 and Figure 2). The prevalence of hypertension increased with age and with BMI category in both sexes, with a steeper increase in prevalence from those aged 21-22 years to 23-24 years, and from the overweight to obesity category, among women than men (Figures 3 and 4). Prevalence was equivalent among the sexes by age 23-24 years (prevalence at 18 years: 4.3% (95% CI:3.5, 5.2) and 7.3% (95% CI:6.2, 8.6), and at 23-24 years 9.50% (95 % CI:8.4, 10.7) and 10.9% (95% CI:9.3, 12.6) among women and men, respectively). The effect of obesity on systolic hypertension was especially marked among men (Supplementary Figure 1), but 6.3% (95% CI:5.8, 6.8) of women were obese compared to only 1.3% (95% CI: 1.0, 1.6) of men. The prevalence of obesity was higher in women at all ages and rose sharply with age, in women aged 23-24 years, compared to those aged 18 years (Supplementary Figure 2).

**Figure 2:**
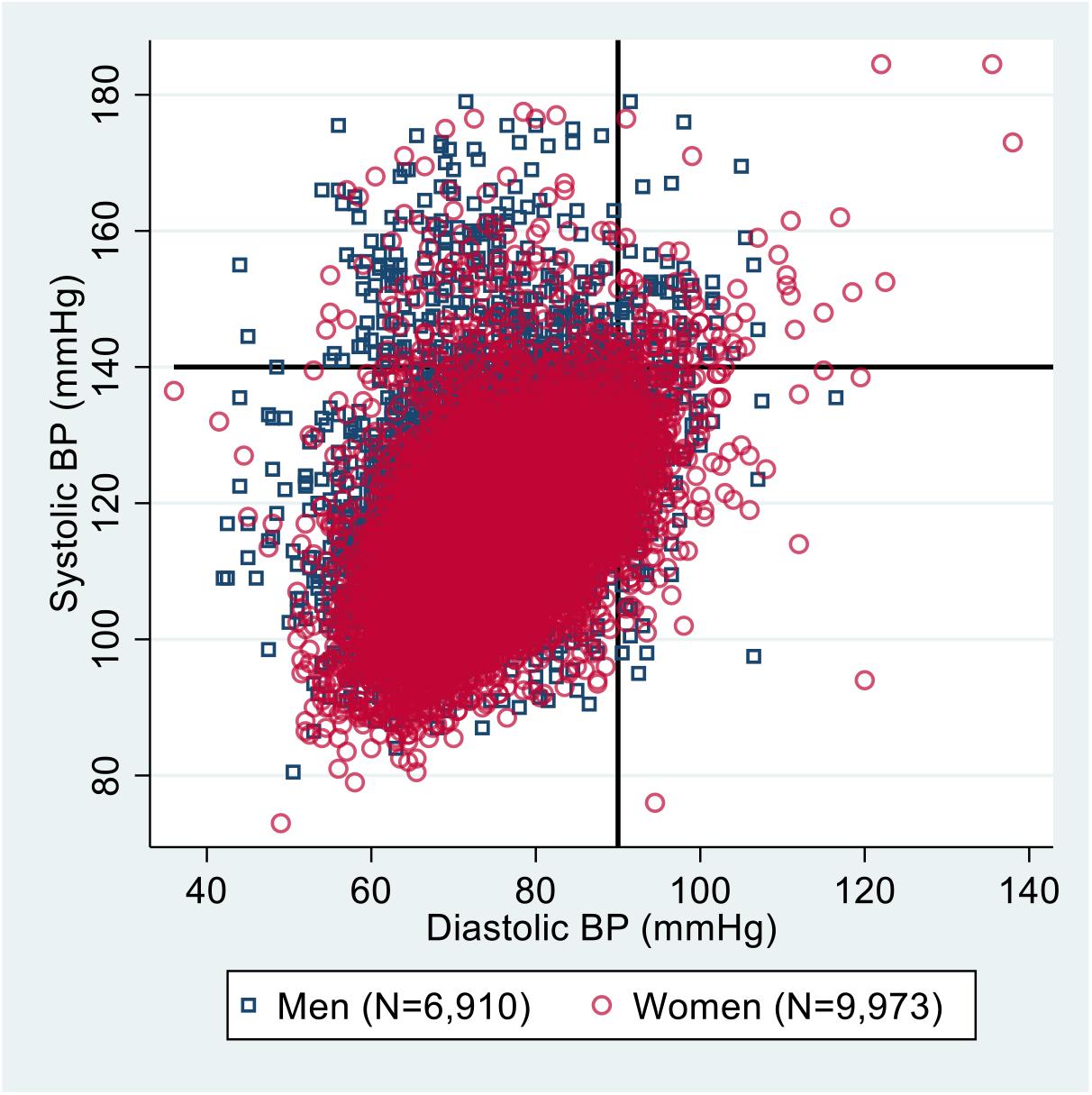
Scatter plot of population BP (excluding pregnant women)

**Figure 3:**
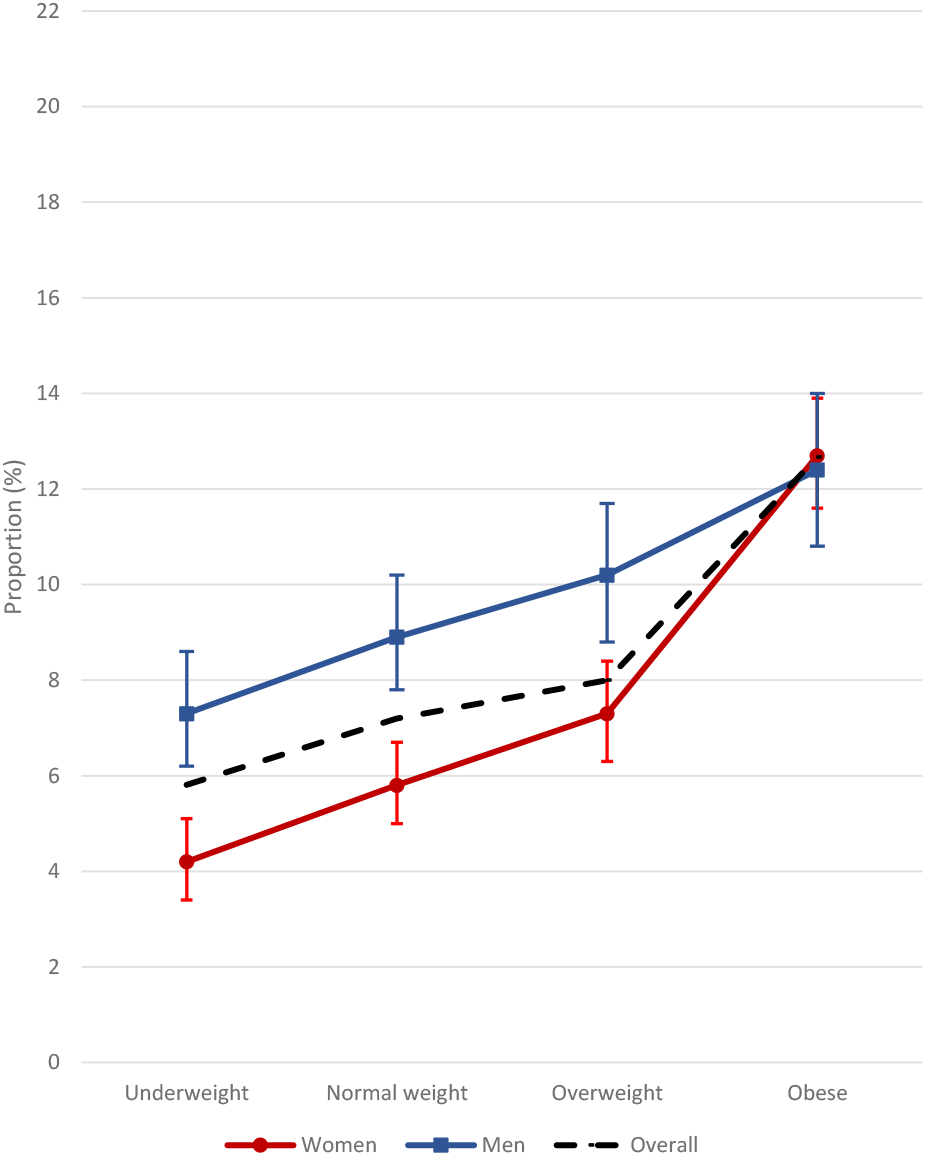
Prevalence of hypertension by age category

**Figure 4:**
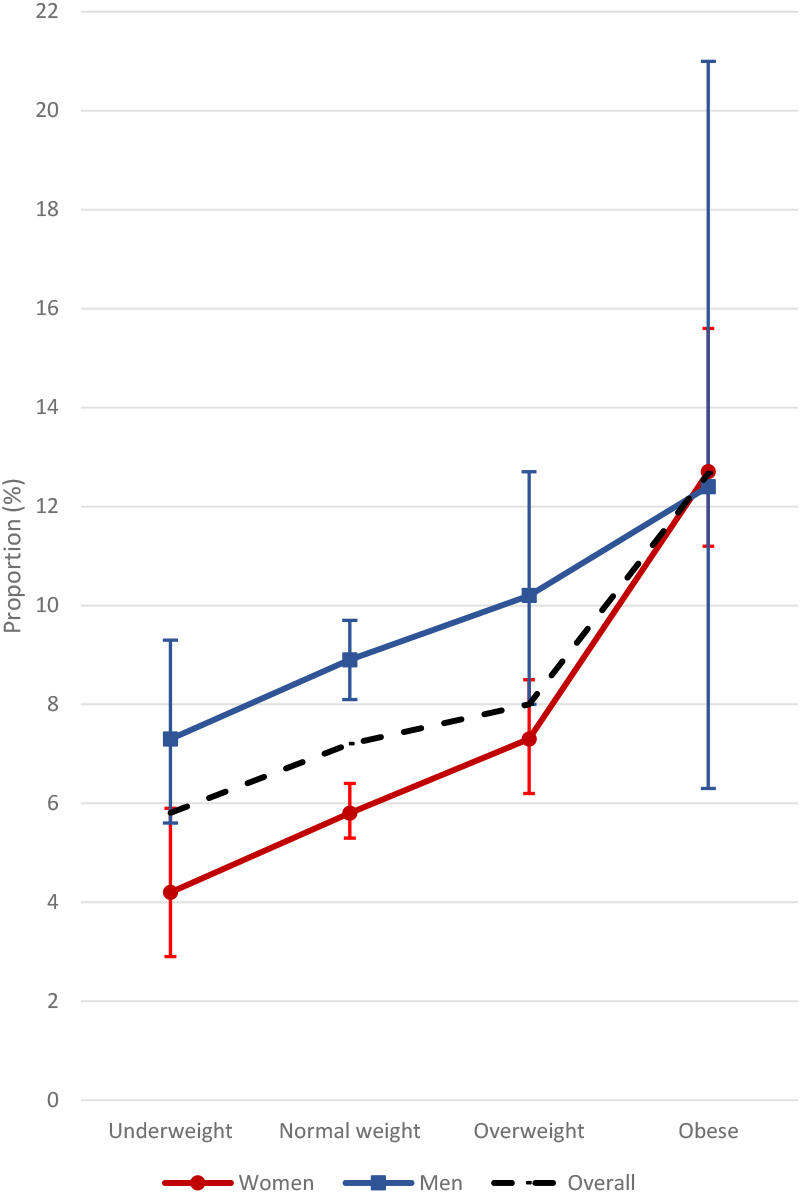
Prevalence of hypertension by BMI category

### Factors associated with hypertension (≥140/90 mmHg)

Male sex, older age, higher level of education, occupation status, socio-economic quintile, higher BMI and HIV status were associated with having hypertension on crude analysis (Table 3). After adjustment for these factors, being male remained strongly associated with hypertension (adjusted Odds Ratio (aOR):1.53; 95% CI:1.36-1.74;); and obesity was the most strongly predictive factor for hypertension (aOR:1.94; 95% CI:1.53-2.47 compared with normal weight). Age was also strongly associated with hypertension, with increasing strength of association with age (19-20 years aOR:1.20; 95%CI:1.00-1.44, 21-22 years aOR:1.45: 95% CI:1.20-1.75 and 23-24 years aOR:1.90; 95% CI:1.57-2.30, compared with 18 years, respectively) (Wald test for trend p<0.001). Being underweight (aOR:0.79; 95% CI:0.63-0.98) and living with HIV (aOR:0.71; 95% CI:0.55-0.92) were associated with lower odds of hypertension. There was insufficient statistical evidence to support interactions between sex and age, or between sex and BMI for the association with hypertension (p=0.09 and p=0.43, respectively). The associations of higher level of education, occupation status and socio-economic quintile with hypertension was no longer apparent in the multivariable model.

**Table 3:**
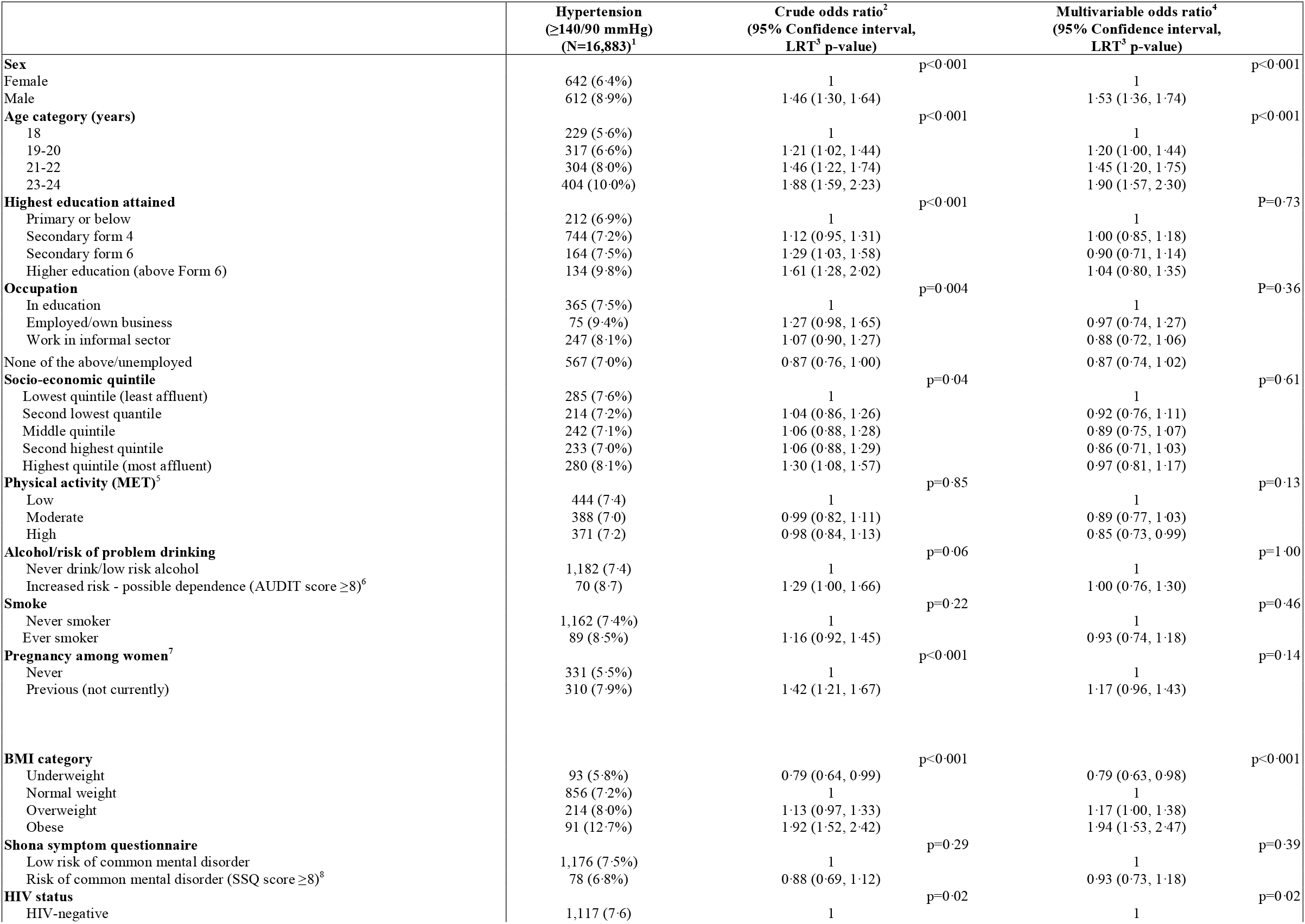
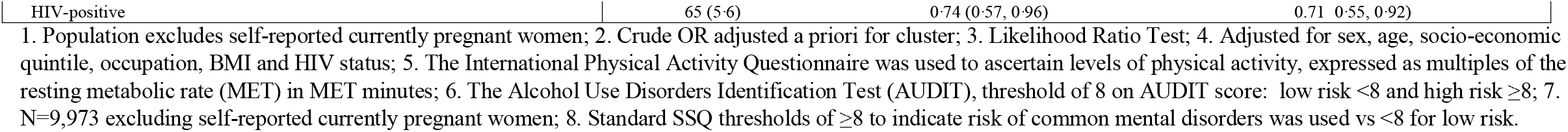
Factors associated with hypertension.

Supplementary Table 2 illustrates that the same risk factors associated with hypertension were also associated with high-normal BP in the multivariable model.

## Discussion

To our knowledge this is the largest population representative survey of BP measures and risk factors for hypertension among youth in SSA, providing much needed robust data on the prevalence of BP elevations in young adulthood. Our study found a high prevalence of hypertension despite the otherwise good health and young age of our population, with 7.4% of young adults aged 18-24 years (9% of men and 7% of women) already meeting criteria for hypertension (≥140/90mmHg). A further 12.2% had high-normal BP (130-139/85-89mmHg) according to ISH guidelines which use more conservative thresholds than current US guidelines (hypertension definition ≥130/80mmHg).^23^ According to the latter 32% of our study sample would be classified as hypertensive.

The prevalence of hypertension (≥140/90mmHg) in our study higher than that from the UK (4% in females and 7% in males aged 16-24 years), despite a much lower prevalence of obesity overall (4% in our study population compared with 12% in 18-24 year olds in the UK).^24,25^ Other limited data involving young adults in SSA have shown similar prevalence of hypertension. In Tanzania, the prevalence of hypertension was 6% of 18-20 year olds and 9% of 21-24 year olds and in Ethiopia prevalence was 10% for 18-25 year olds.^12,26^ Both studies involved about 600 young adults. Our data are consistent with these estimates, but the large scale and population random sampling in our study provide assurance about the precision and representativeness of the estimates.

We observed a sharp rise in hypertension with age, from 6% among 18 year olds to 10% by age 23-24 years. Reasons for the high prevalence and earlier onset of hypertension in SSA include transitions from healthy traditional lifestyles (e.g. diet and physical activity) to unhealthy alternatives including consumption of processed foods high in salt, fat and sugar, alcohol excess, lower physical activity and exposure to air pollution.^18^ Lifetime exposure to socio-demographic pressure which begins in utero, for instance with maternal malnutrition, may contribute to sympathetic activation and early vascular ageing; furthermore, a genetic predisposition causes abnormal sodium homeostasis.^5^ A long-term US follow-up study found that by age 50 years, among individuals with onset of hypertension (≥140/90) at <35 years, 60% had single organ damage, and 25% had concurrent damage in more than one organ.^27^ Our data suggest that young adults in Zimbabwe may be experiencing harmful arterial pressures which will accumulate with tim and could have substantial population impact.

Hypertension prevalence was higher among young men, driven by systolic rather than diastolic elevation. The clinical relevance of isolated systolic hypertension in youth is debated. Growing evidence suggests that it is associated with excess cardiovascular risk at older age, and regular monitoring and lifestyle modification is advised.^28^ In both males and females, BP increased from 18 years to 23-24 years, with hypertension in females equivalent to that among men by age 24 years, likely explained by higher levels of obesity in females.

Being obese was associated with almost twice the odds of having hypertension, consistent with other studies across age groups.^2,13^ Critically, the global prevalence of obesity has tripled over the last 50 years and in SSA, undernutrition and obesity co-exist as public health challenges.^29^ In our study, a fifth of youth were overweight or obese but a further 9% were underweight. In many cultures, higher weight is traditionally perceived to be associated with better health and therefore nuanced, culturally sensitive messaging is required to address over- and under-nutrition. The prevalence of obesity in our study increased from 3% to 8% between the ages of 18 years to 23-24 years, largely driven by the increase in prevalence among women. Overweight was also highly prevalent among women, and this can be expected to increase with advancing age or evolve into obesity. Consequently, the prevalence of hypertension in women as they age may even exceed that in men as seen in other studies on older Africans.^30^

Consistent with other studies from SSA, people living with HIV were less likely to have hypertension even after adjusting for BMI.^31^ This is in contrast to findings from HICs where HIV increases cardiometabolic risk. Possible explanations include race differences and socio-demographic factors;^5^ however, further research is warranted. While we did not find evidence to support an association between common mental disorders and hypertension, further exploration of the effect of chronic stress on hypertension remains relevant.

A question that arises in studies which measure BP at a single time point is whether these findings may represent an artificially elevated blood pressure as a result of “white coat” hypertension and whether a measure on a single occasion represent persistently elevated BP. The clear age-trends in our study, and association of hypertension with known risk factors consistent with other studies, suggest our findings are unlikely to be spurious. The high prevalence of high-normal BP and the finding that risk factors for high-normal BP were the same as for hypertension, illustrate that elevated BP measures represent a continuum. Thus, our findings highlight an urgent and neglected public health problem in Africa, with the consequent morbidity and mortality only likely to be visible in a decade. Intervening in this age-group represents a timely opportunity to avert the considerable mortality and morbidity due to cardiovascular disease that is being observed among older adults, now the leading cause of death globally.

Further research to investigate BP in SSA youth is urgently needed. Specifically, unattended BP measures, and 24-hour ambulatory BP monitoring would enable interrogation of both the possible “white-coat” effect and of masked hypertension, a term which refers to the lack of normal lowering of BP at night time (non-dipping).^6^ In addtion, it is vital to interrogate the aetiological factors for development of hypertension. The advances in technologies to measure blood pressure, including non-invasive techniques for central blood pressure monitoring and organ imaging, and the advent of metabolomics, offer opportunities to interrogate whether these blood pressure measurements are “real” and persistent and are associated with metabolic perturbations, and whether there is end-organ damage, well before cardiovascular events occur.^32,33^ The optimal timing to recommend drug treatment is a further complex context specific question which has to be examined and weighed up, considering realities of health systems in SSA against benefits to individual health.

A key strength of this study was that it had a very large sample size, with a population representative sample of urban and peri-urban youth. The survey was conducted across a large number of peri-urban and urban communities across the country and participation rates were high. Multiple risk behaviours and health outcomes were assessed, and validated data collection tools, international guidelines for BP measurements and definitions were used. A limitation of our study was that BP was measured during a single visit. However, other internationally accepted standards were adhered to with three seated measurements taken at standardised intervals and the average of the second and third measures were used to determine hypertension, an approach supported by our finding that the first measure was consistently higher than the latter two measures.^7^Assessment of other traditional risk factors for cardiovascular disease such as dyslipidaemia and hyperglycaemia was not undertaken. Behaviour data e.g. alchol consumption or physical activity were self-reported and may be subject to recall or desirability bias but it is unlikely that it was differential by BP measure.

In summary, our study demonstrates that hypertension is already prevalent at an alarming level among youth in urban and peri-urban settings in Zimbabwe. Further research to understand the drivers of hypertension, impact on target organs, as well as sustainable lifestyle modification strategies and indications for treatment initiation in youth in SSA, is urgently needed.

## Data Availability

Individual, anonymized participant data and a data dictionary will be available through the LSHTM repository (Data Compass) 12 months after publication of trial results. Data will be available to anyone for further analyses with approval from the Medical Research Council of Zimbabwe.

## Contributors

RAF and RJH designed the study. RAF, ED and CDC coordinated the study. CDC, KK and ED contributed to the study design and study logistics. TB and VS were responsible for data management. KS analysed the data with input from FCM, VS and RAF. DR, AS, AD, AES provided expert advice on study tools. All authors contributed to writing the report and have seen and approved the final draft.

## Declaration of interests

RAF’s institution received a grant from the Wellcome Trust. Salary support for VS and RH was in part from a grant from the Medical Research Council (MRC) and the Department for International Development (DFID UK) under the MRC/DFID Concordat (MR/K012126/1). All other authors declare no competing interests.

## Acknowledgements

The study was funded by the Wellcome Trust (Grant 095878/Z/11/Z). We thank all participants and field teams.

## Data sharing

Individual, anonymized participant data and a data dictionary will be available through the LSHTM repository (Data Compass) 12 months after publication of trial results. Data will be available to anyone for further analyses for with approval from the Medical Research Council of Zimbabwe.

## Notes

### Author Declarations

The study was approved by the Medical Research Council, Zimbabwe, the Biomedical Research and Training Institute Institutional Review Board and the ethics committee of the London School of Hygiene and Tropical Medicine. Participants viewed an information video about the study (in either English, Shona or Ndebele) on a tablet. Consent was documented electronically on a tablet, with participants retaining a signed paper copy for their records.

