## Supplementary Material for "High prevalence of hypertension and high-normal blood pressure: findings from a large population-based survey of young adults in Zimbabwe"

### Supplementary Table 1: Median BP measures at first, second and third reading

| **BP measurements** N=16,883 | **Median systolic BP (IQR) mmHg** | **Median diastolic BP (IQR) mmHg** |
|---|---|---|
| 1st measurement | 118 (110, 126) | 75 (69, 81) |
| 2nd measurement | 116 (109, 125) | 74 (69, 80) |
| 3rd measurement | 116 (109, 124) | 74 (68, 80) |

**Supplementary Table 2: Factors associated with high-normal BP**

| | High-normal (Systolic 130-139/ Diastolic 85-89 mmHg) (N=15,629)[1] | Crude odds ratio[2] (95% Confidence interval, LRT[3] p-value) | Multivariable odds ratio[4] (95% Confidence interval, LRT[3] p-value) |
|---|---|---|---|
| **Sex** | | p<0·001 | p<0·001 |
| Female | 1033 (11·1%) | 1 | 1 |
| Male | 1031 (16·4%) | 1·63 (1·48, 1·79) | 1·70 (1·54, 1·89) |
| **Age category (years)** | | p<0·001 | p<0·001 |
| 18 | 426 (11·0%) | 1 | 1 |
| 19-20 | 521 (11·6%) | 1·04 (0·91, 1·20) | 1·01 (0·88, 1·17) |
| 21-22 | 515 (14·2%) | 1·33 (1·16, 1·53) | 1·28 (1·11, 1·49) |
| 23-24 | 602 (16·6%) | 1·58 (1·38, 1·81) | 1·52 (1·31, 1·77) |
| **Highest education attained** | | p=0·005 | p=0·16 |
| Primary or below | 379 (13·3%) | 1 | 1 |
| Secondary form 4 | 1210 (12·7%) | 0·97 (0·85, 1·10) | 0·90 (0·80, 1·03) |
| Secondary form 6 | 270 (13·4%) | 1·05 (0·89, 1·25) | 0·93 (0·78, 1·11) |
| Higher education (above Form 6) | 205 (16·6%) | 1·32 (1·10, 1·60) | 1·08 (0·87, 1·32) |
| **Occupation** | | p<0·001 | p=0·25 |
| In education | 576 (12·7%) | 1 | 1 |
| Employed/own business | 101 (13·9%) | 1·09 (0·87, 1·37) | 0·93 (0·74, 1·18) |
| Work in informal sector | 435 (15·5%) | 1·25 (1·09, 1·43) | 1·14 (0·98, 1·33) |
| None of the above | 952 (12·6%) | 0·97 (0·87, 1·09) | 1·06 (0·93, 1·20) |
| **Socio-economic quintile** | | p=0·17 | p=0·48 |
| Lowest quintile (least affluent) | 468 (13·6%) | 1 | 1 |
| Second lowest quantile | 367 (13·3%) | 1·02 (0·88, 1·19) | 0·98 (0·85, 1·14) |
| Middle quintile | 403 (12·8%) | 1·01 (0·87, 1·18) | 0·92 (0·79, 1·07) |
| Second highest quintile | 382 (12·3%) | 0·98 (0·84, 1·15) | 0·89 (0·76, 1·03) |
| Highest quintile (most affluent) | 444 (14·1%) | 1·16 (1·00, 1·36) | 0·98 (0·84, 1·14) |
| **Physical activity (MET)**[5] | | p=0·02 | p=0·28 |
| Low | 662 (12·0) | 1 | 1 |
| Moderate | 699 (13·6) | 1·14 (1·02, 1·28) | 1·09 (0·97, 1·22) |
| High | 650 (13·6) | 1·16 (1·03, 1·31) | 1·02 (0·90, 1·14) |
| **Alcohol/risk of problem drinking** | | p=0·26 | p=0·11 |
| Never drink/low risk alcohol | 1,960 (13·2) | 1 | 1 |
| Increased risk - possible dependence (AUDIT score ≥8)[6] | 102 (14·0) | 1·13 (0·91, 1·41) | 0·83 (0·66, 1·05) |
| **Smoke** | | P=0·002 | P=0·74 |
| Never smoker | 1908 (13·0%) | 1 | 1 |
| Ever smoker | 156 (16·2%) | 1·35 (1·13, 1·61) | 0·97 (0·80, 1·17) |
| **Pregnancy among** women[7] | | p<0·001 | p<0·40 |
| Never | 573 (10·0%) | 1 | 1 |
| Previous (not currently) | 460 (12·8%) | 1·29 (1·13, 1·47) | 1·07 (0·91, 1·26) |
| **Shona symptom questionnaire** | | p=0·10 | p=0·45 |
| Low risk of common mental disorder | 1,939 (13·3%) | 1 | 1 |
| Risk of common mental disorder (SSQ score ≥8)[8] | 125 (11·7%) | 0·85 (0·70, 1·03) | 0·93 (0·76, 1·13) |
| **BMI category** | | p<0·001 | p<0·001 |
| Underweight | 154 (10·2%) | 0·80 (0·67, 0·95) | 0·77 (0·64, 0·92) |
| Normal weight | 1422 (12·9%) | 1 | 1 |
| Overweight | 382 (15·5%) | 1·24 (1·10, 1·41) | 1·35 (1·19, 1·54) |
| Obese | 106 (16·9%) | 1·34 (1·08, 1·67) | 1·55 (1·24, 1·94) |
| **HIV status** | | p=0·02 | p=0·02 |
| HIV-negative | 1,1936 (13·4) | 1 | 1 |
| HIV-positive | 120 (10·9) | 0·80 (0·66, 0·97) | 0.80 0·66, 0·98) |

Excludes participants with hypertension (n=1,254); 2. Crude OR adjusted a priori for cluster; 3. Likelihood Ratio Test; 4. Adjusted for sex, age, socio-economic quintile, occupation, BMI and HIV status; 5. The International Physical Activity Questionnaire was used to ascertain levels of physical activity, expressed as multiples of the resting metabolic rate (MET) in MET minutes; 6. The Alcohol Use Disorders Identification Test (AUDIT), threshold of 8 on AUDIT score: low risk <8 and high risk ≥8; 7. N=9331 excluding self-reported currently pregnant women; 8. Standard SSQ thresholds of ≥8 to indicate risk of common mental disorders was used vs <8 for low risk.

**Supplementary Figure 1: Prevalence of systolic hypertension (any diastolic BP) by BMI category**

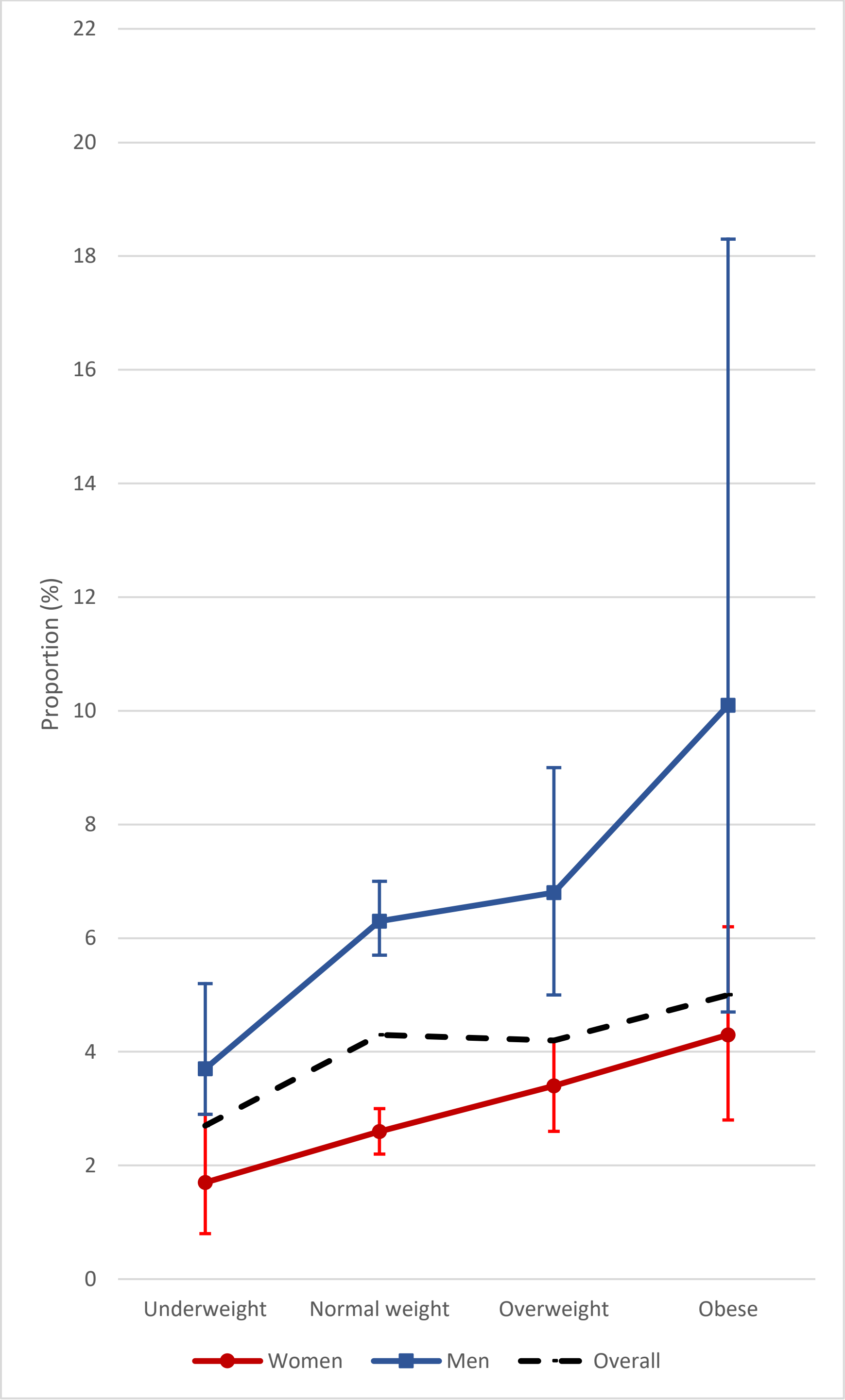

**Supplementary Figure 2: Prevalence of obesity with age**

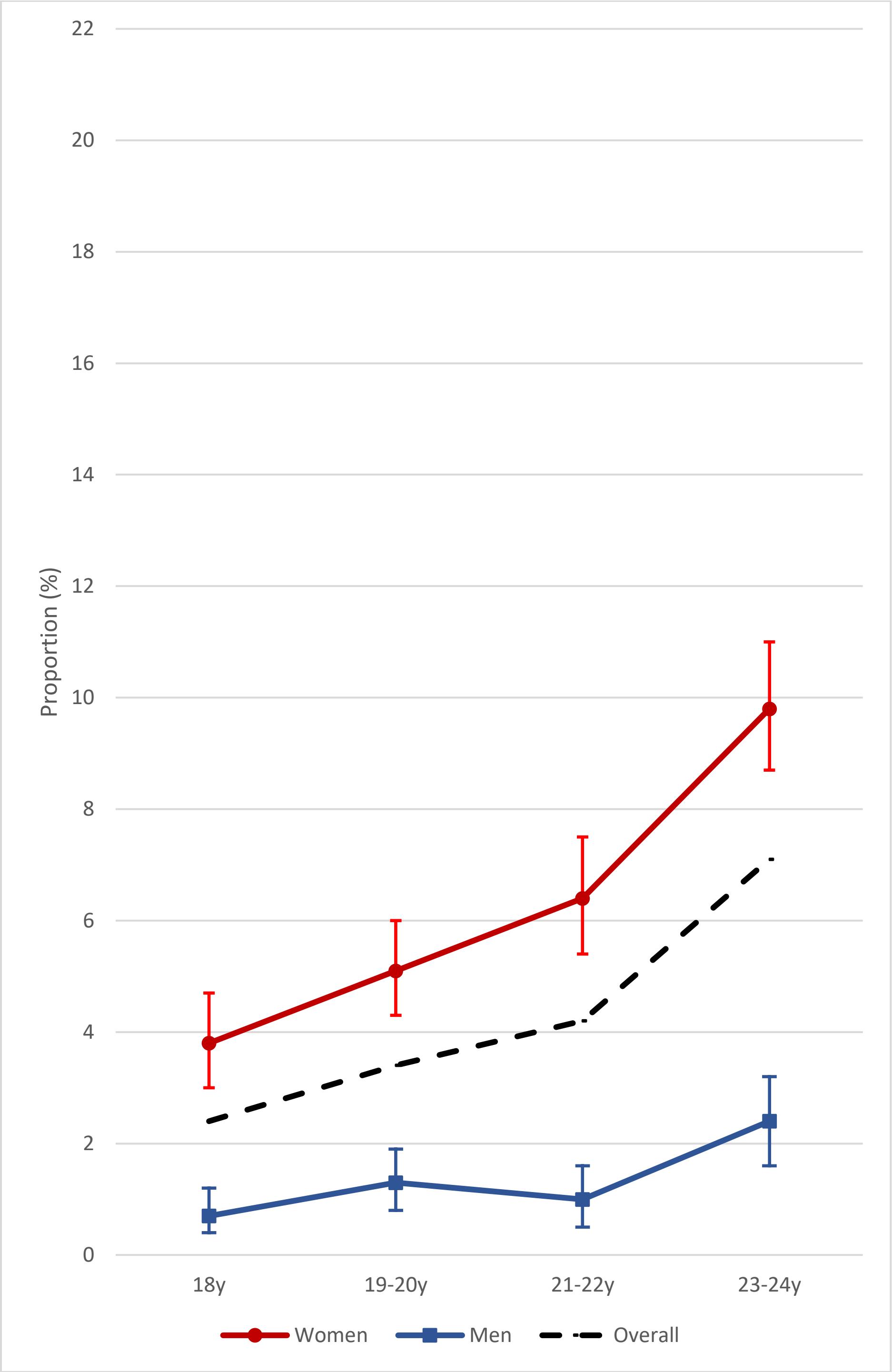
